# Persistence of SARS-CoV-2 antibodies and symptoms in an Irish Healthcare Worker (HCW) setting: Results of the COVID Antibody Staff Testing (CAST) Study

**DOI:** 10.1101/2021.02.10.20248323

**Authors:** Joanna Griffin, Elizabeth Tully, Fiona Cody, Katherine Edwards, Kara Moran, David LeBlanc, John O’Loughlin, Ruth McLoughlin, Rachel Cummins, Sheila Breen, Richard Drew, Dominick Natin, Fergal Malone

## Abstract

**Objectives:** This study examined the natural history, including incidence and prevalence, of SARS-CoV-2 antibodies serially up to 6 months post infection in Irish Healthcare Workers (HCWs) at an academic tertiary maternity hospital, during the first pandemic peak from March to September 2020.

**Design:** This single centre observational study profiled SARS-CoV-2 incidence and infection using viral RNA detected using oro/nasopharyngeal swabs accompanied by serological assessment of study participants for the presence of S SARS-CoV-2 antibodies. Participant demographics were also collected alongside information on clinical symptoms and time to recovery. Real time polymerase-chain-reaction (RT-PCR) for viral RNA SARS-CoV-2 detection was performed using the Allplex™ SARS-CoV-2 three gene target 2019-nCoV assay (SeeGene Inc., Rep. of Korea) or the Xpert Xpress SARS-CoV-2 assay on the GeneXpert platform (Cepheid, USA). Blood samples were obtained at the time of initial swab and at up to 4 time points thereafter, for the serological assessment of antibodies against both the spike and nucleocapsid protein antigens of SARS-CoV-2. Serological response was measured using the Captia™ Anti-SARS-CoV-2 (IgG) ELISA (Trinity Biotech) as part of a clinical performance evaluation. Two other testing methods were also used; the Anti-SARS-CoV-2 ELISA (IgG) assay (EuroImmun) and the Abbott Anti-SARS-CoV-2 IgG 75 kit on the Architect™ i2000SR instrument (Abbott Laboratories).

**Setting:** Academic Tertiary Maternity Hospital in Dublin, Ireland.

**Participants:** We invited symptomatic and asymptomatic healthcare workers employed at the Rotunda Maternity Hospital to participate in the CAST study.

**Main Outcome Measures:** The CAST study aimed to examine incidence and clinical symptoms of SARS-CoV-2 in HCWs and to determine the presence and longevity of antibodies in this group. We also sought to examine the clinical utility of the Captia™ Anti-SARS-CoV-2 (IgG) ELISA (Trinity Biotech) and to compare it to the current “accepted” gold standard platform in Ireland.

**Results:** By July 2020, 398 molecular tests had been completed on symptomatic staff with clinical suspicion of SARS-CoV-2 infection. In this cohort, 14% (n=54/398) had SARS-CoV-2 RNA detected by RT-PCR. The CAST study enrolled 137 HCWs with 86 participants symptomatic at time of swab collection and a further 51 asymptomatic control participants. SARS-CoV-2 RNA was detected in 52% (n=45/86) symptomatic study participants and serological positivity was confirmed in 98% (n=44/45) of those participants. Asymptomatic SARS-CoV-2 RNA infection was detected in 4% (n=2/51) of control participants with a seropositivity rate in this group of 8% (n=4/51). We demonstrated that 95% of SARS-CoV-2 PCR positive participants have detectable levels of antibodies at 100 days post infection, which persisted in 91% of participants at day 160+. Ongoing symptoms up to six months post infection were present in 50% of study participants with positive PCR and serology results. These data will be important to consider for long-term workforce planning in a healthcare setting, as the ongoing pandemic continues.

**Funding:** The CAST study was supported by the Rotunda Hospital and Trinity Biotech.

## Introduction

The SARS-CoV-2 pandemic reached the Republic of Ireland in February 2020 and by March 27^th^, Ireland, along with many other countries, introduced a lockdown to inhibit the spread of the virus.^1^ HCWs, in maintaining healthcare services and clinical activity continued to be exposed to patients and other staff and are therefore considered to be a high risk cohort for contracting SARS-CoV-2.^2,3,4^ HCWs currently account for approximately 25% of the infected population in Ireland with information on the longer term effects of SARS-CoV-2 infection in this cohort continuing to emerge. ^5,6,7^

Robust testing strategies have shown to be an important tool in slowing down the pandemic, supporting decisions on infection control and improving the knowledge of virological aspects of SARS-CoV-2.^8,9^ However, testing should be accompanied by clear standardized comprehensible information on performance for both clinicians and patients. The European Centre for Disease Control (ECDC) and the World Health Organisation (WHO) currently recommend COVID-19 diagnosis via the identification of viral genome targets by RT-PCR in respiratory tract materials during the first week of symptoms for SARS-CoV-2 virus RNA.^8,9^ This type of test is most useful when the virus is actively replicating, therefore identifying acute or early infection, with sensitivity ranging from 34% to 80%.^8^ While it is acknowledged that molecular testing may yield false positive and negative results, it is still the most common and reliable test across Europe. ^10,11,12^

Serology testing which determines the presence or levels of antibodies to SARS-CoV-2 in the blood is useful in identifying people who have developed immunity to the virus. Detection of IgM antibodies can be found in serology samples from one week post-infection with IgG antibodies or long term immunoglobulins detectable from Days 7-21.^13,14^ The longevity of detected antibodies and whether this infers protective immunity is still unclear with information on future defence against COVID-19 continuing to evolve as ongoing research emerges.^15^

The need for a locally manufactured, sensitive and robust open platform diagnostic system is important for the ongoing fight against COVID-19. Trinity Biotech has developed an Enzyme-linked Immunosorbent Assay (ELISA), Captia™ SARS-CoV-2 IgG kit for the qualitative detection of human IgG class antibodies to SARS-CoV-2 in human serum. The Captia™ SARS-CoV-2 IgG kit is an *in vitro* diagnostic (IVD) device intended for use in a laboratory setting. The CAST study is a collaborative project between Trinity Biotech and the Rotunda Hospital which sought to examine the incidence of SARS-CoV-2 in HCWs during the first pandemic peak.

## Methods

This single centre prospective observational study was conducted at the Rotunda Hospital in Dublin from late March to September 2020. The study was approved by the Hospital Research Ethics Committee (REC-2020-012). Hospital Staff were invited to participate by the Occupational Health Department at a COVID-19 pre-swabbing consultation, with asymptomatic controls invited to volunteer via a hospital-based poster and email campaign. All tests were carried out in a designated staff testing area of the hospital, with same day PCR results available. An on-site contact tracing team was available to quickly remove close contacts of positive staff in order to curb transmission. Participants provided written informed consent for collection of both oro/nasopharyngeal swabs and serum samples. Participants were informed of their swab result within 12 hours of sample collection, with serum samples batched, stored at −20°C and subsequently tested at the end of the study period. Serial follow-up serum samples were obtained from study participants and a second swab sample taken from study controls at the time of last blood draw to ensure continued absence of disease. Additional eligibility criteria for enrolment in the CAST study included staff being currently employed at the hospital, those fluent in English, and those aged ≥18 years. All participants were counselled appropriately and written informed consent was obtained. Demographic data including age, gender, ethnicity, medical history as well as presenting and ongoing symptoms were collected on all participants and transferred to a confidential secure database for analysis.

### Statistical Analyses

At study design stage, it was estimated that the asymptomatic prevalence of SARS-CoV-2 would likely range between 30-40%.^16^ A recent Spanish study of HCWs found a cumulative prevalence of 11.2%, with seroprevalence in the general Irish population estimated at 1.7% and HCW seroprevalence in Ireland reported to be closer to 18%.^17,18,19^

### Testing Strategy

Molecular Testing for SARS-CoV-2 infection was initiated on March 16^th^ 2020 and by the end of July 2020, 398 PCR tests had been completed via the Occupational Health Department on symptomatic staff with clinical suspicion of SARS-CoV-2 infection. From May 12^th^ 2020, the addition of serological screening was introduced for both symptomatic and asymptomatic staff at the hospital with a previous or current laboratory-confirmed swab result for SARS-CoV-2 infection. Serial blood samples (n=598) were collected on 137 participants who had either an oro/nasopharyngeal swab taken at study enrolment (n=92) to determine current infection or a laboratory-confirmed positive swab result (n=45) within the last 6 weeks. Serial blood draws were obtained every 2-3 weeks for prospective participants with recovered participants volunteering three samples at approximately day 50, 100 and 160+ post-confirmation of COVID-19 infection. Control participants had a laboratory-confirmed negative PCR swab at enrolment and again at the time of last blood draw, in order to confirm ongoing absence of disease. Figure 1 shows the CAST study flow chart.

**Figure 1.**
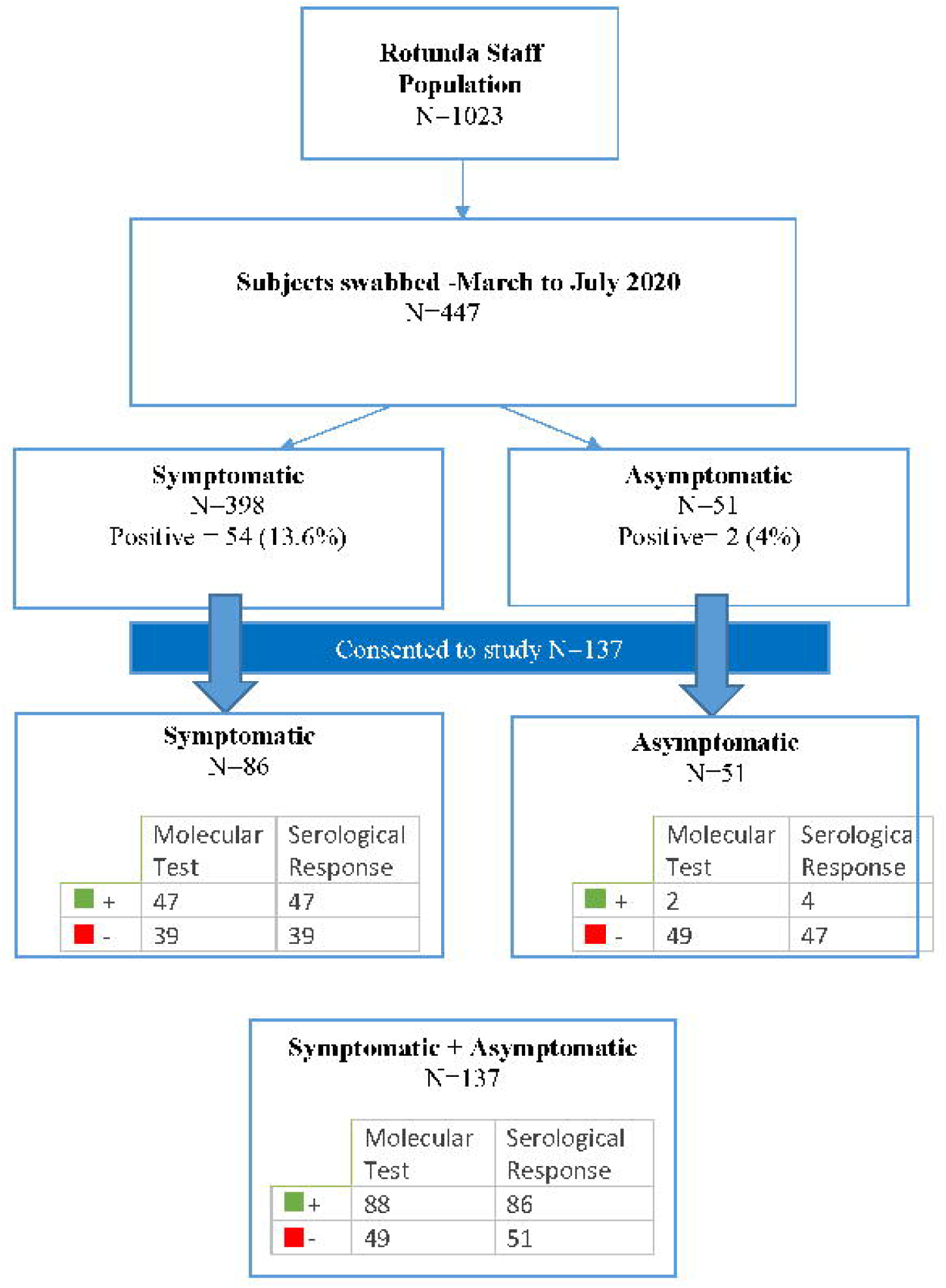
CAST Study Flow.

### Viral RNA Detection

SARS-CoV-2 RNA testing was performed on either the Allplex™ 2019-nCoV assay (manufactured by SeeGene Inc, Rep. of Korea) or Xpert^®^ Xpress SARS-CoV-2 (manufactured by Cepheid, USA) in accordance with the manufacturer’s instructions. Results were reported as either “detected” (targets 1 and 2 detected), “presumptive positive” (target 1 not detected; target 2 detected), or “not detected”.

### Serology

Serum samples were collected from participants, processed immediately and stored at −20°C prior to transport to Trinity Biotech, Bray, Ireland for analysis. Samples were tested using the Captia™ SARS-CoV-2 IgG assay following the manufacturer’s instructions. The assay utilises ELISA technology for the qualitative detection of IgG in human serum against the SARS-CoV-2 spike protein 1 (S1) and 2 (S2) subunits. The Captia™ requires a volume of 10μL of serum for the detection of IgG antibody to SARS-CoV-2 (10μL diluted with 200μL of sample diluent). Qualitative results were calculated using the absorbance readings generated by the samples and kit controls. Serum samples which were discordant with their corresponding molecular PCR result were also tested using an alternative CE marked kit: the anti-SARS-CoV-2 ELISA (IgG) assay (EUROIMMUN). This assay also utilises ELISA technology for the qualitative detection of IgG in human serum or plasma against the SARS-CoV-2 S1 subunit. The anti-SARS-CoV-2 ELISA (IgG) requires a sample volume of 10μL for the detection of IgG antibody to SARS-CoV-2 (10μL diluted with 1mL of sample diluent). Qualitative results were calculated using the absorbance readings generated by the samples and kit controls.

In addition, serum samples from all 137 study participants were tested on the Abbott Architect™ i2000SR instrument using the Abbott SARS-CoV-2 IgG75 assay following the manufacturer’s instructions. This additional method was selected as it is the current chosen assay used by the Health Service Executive for seroprevalence assessment in Ireland. This testing took place in the clinical diagnostic laboratory at the Rotunda Hospital. The assay is a two-step immunoassay using chemiluminescent microparticle technology for qualitative detection of IgG in human serum or plasma against the SARS-CoV-2 nucleoprotein. The Abbott Architec™t assay requires a volume of 75µL of serum or plasma for the detection of IgG antibody to SARS-CoV-2 (25µL test volume and 50µL dead volume). Qualitative results and index values reported by the instrument were used for analysis.

## Results

Of the 1,023 staff at the Rotunda hospital, 44% or 449 (398 symptomatic and 51 asymptomatic controls) individuals were tested between March 16^th^ and July 31^st^ 2020 (Figure 2). In the symptomatic group, 14% (n=54/398) had detectable levels of SARS-CoV-2 RNA in their swab sample. Of the 54 positive staff members during this timeframe, 49 consented to participate in the CAST study. The CAST study enrolled 137 participants in total (as per Figure 1) and oro/nasopharyngeal RT-PCR results were available for all 137 participants. Five hundred and ninety eight individual serum samples were collected on these participants, and serological positivity for SARS-CoV-2 was confirmed in 98% (48/49) of PCR positive participants. Study demographics are presented in Table 1, whereby, the median age of participants is reported as 42 years with almost 90% of participants reporting no co-morbidity. Less than 32% had close contact with a confirmed SARS-CoV-2 infection. The first peak of infection of staff in the Rotunda was on 25^th^ March 2020 when 6 staff members tested positive.

**Table 1:** CAST Study Demographics.

|  |  |
| --- | --- |
| **Age** | **N=137 (100%)** |
| Median Age (years) | 42 |
| **Ethnicity** |  |
| White (%) | N=120 (88%) |
| Black/Black Irish (%) | N=3 (2%) |
| Asian/Asian Irish (%) | N=14 (10%) |
| Other (including mixed background) (%) | N=0 (0%) |
| **Co Morbidities** |  |
| Yes (%) | N=15 (11%) |
| No (%) | N=122 (89%) |
| **Close contact with confirmed SARS-CoV-2 case** |  |
| Yes (%) | N=44 (32%) |
| No (%) | N=82 (60%) |
| Unknown (%) | N=11 (8%) |
| **Gender** |  |
| Male | N=16 (12%) |
| Female | N=121 (88%) |

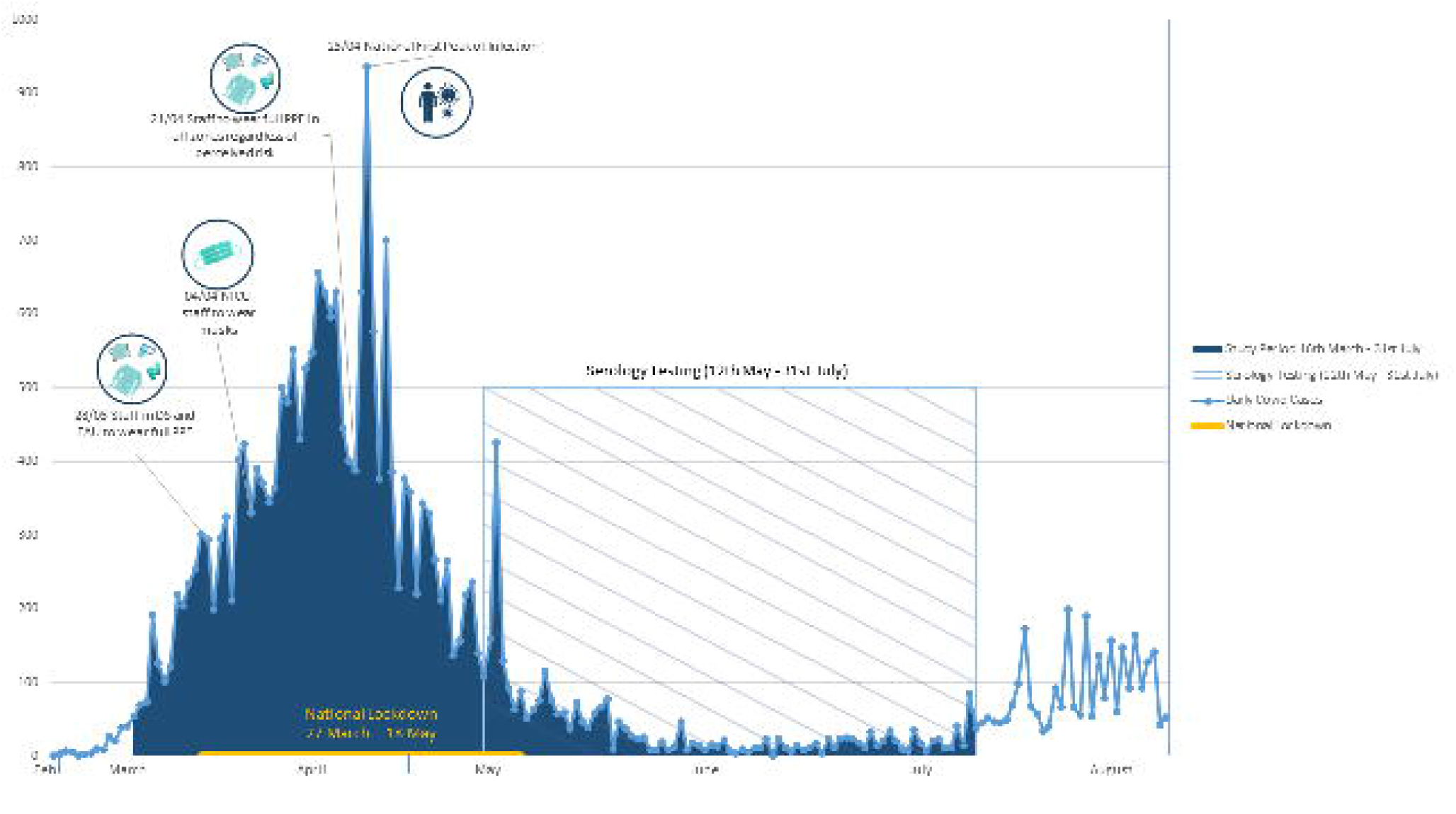

Asymptomatic SARS-CoV-2 viral RNA infection was detected in 4% (2/51) control participants, with a seropositivity rate in this group of 8% (4/51 asymptomatic seropositive participants). Table 2 shows testing data for participants with both confirmed SARS-CoV2 infection via PCR and the detection of SARS-CoV-2 antibodies at Day 50 (using both the Captia™ and Abbott testing methods). All recovered study participants were approached for a blood sample up to 6 months post initial infection and samples were obtained from 45 of these participants. Analysis of these samples was performed by Trinity Biotech using the Captia™ assay, as before. Results showed that 95% of recovered participants had detectable antibody in their serum at day 100 with 91% (41/45) continuing to have detectable anti-SARS-CoV2 antibody in their serum after day 160. Only 9% (4/45) of participants exhibited a decrease in detectable levels of antibodies at 6 months post exposure to Covid-19. Data from the CAST study was used to support a performance claim for the Captia™ SARS-CoV-2 IgG ELISA for Positive Percent Agreement (PPA) and Negative Percent Agreement (NPA) at ≥14 days from day 0 (date of oro/nasopharyngeal swab) in a European population.

**Table 2:** CAST Study Results Correlation of Molecular & Serological Testing.

| Molecular PCR Test | Serological Test |  |  |  |
| --- | --- | --- | --- | --- |
|  | Captia™ SARS-CoV-2 IgG assay |  | Abbott™ SARS-CoV-2 IgG75 |  |
| Total N=137 | Detected | Non Detected | Detected | Non Detected |
| ***Symptomatic HCWs (N=86)*** |  |  |  |  |
| Positive (N=47) | 46/47 (98%) | 1/47 (2%) | 38/47 (81%) | 9/47 (19%) |
| Negative (N=39) | 0/39 (0%) | 39/39 (100%) | 0/39 (0%) | 39/39 (100%) |
| ***Asymptomatic HCWs (N=51)*** |  |  |  |  |
| Positive (N=2) | 2/2 (100%) | 0/2 (0%) | 2/2 (100%) | 0/2 (0%) |
| Negative (N=49) | 0/49 (100%) | 49/49 (100%) | 0/49 (100%) | 49/49 (100%) |

Consent was obtained from all study participants to allow access to Occupational Health Covid-19 records and medical history which included presenting, day 5 and day 12 symptoms. The most common presenting symptoms for SARS-CoV-2 positive staff were cough (67%), sore throat (42%) and fever (37%). At day 5 post positive PCR result, the most commonly reported symptom remained a cough (45%), with shortness of breath (40%) and fever (38%) featuring prominently. By day 12 fatigue (36%), cough (32%) and shortness of breath (21%) were most commonly reported, with just over 23% reporting being symptom-free at this point. Approximately 68% of symptomatic RT-PCR positive participants did not need any further medical assistance with 30% requiring assessment by a GP, 9% needing medical assessment in hospital, and one individual requiring hospitalisation for further management of symptoms. For SARS-CoV-2 positive subjects the average duration of time absent from work was 21 days from onset of symptoms with the average time to recovery increasing with increasing age. Staff members over the age of 50 were absent for 5 more days on average, compared with younger staff aged <50 years.

All recovered staff who previously had a confirmed positive PCR test were contacted in mid-September and surveyed with regard to ongoing symptoms. In this group, 50% of PCR positive participants reported having ongoing symptoms and of this group 60% reported ongoing fatigue, with 28% experiencing continued shortness of breath and a further 20% reporting ongoing loss of taste and/or smell. Of note, seven have been prescribed a corticosteroid inhaler for ongoing management of symptoms.

## Discussion

Molecular and serological assays for SARS-CoV-2 are being developed and deployed at a rapid rate. While some serological tests obtained rapid approval during the early phases of the pandemic, the quality and robustness of many testing methods has been questioned, with lessons having been learned about high levels of false positive and false negative results and the implications of this.^20,21^ Our experience with SARS-CoV-2 highlights the need for a common legislative framework to ensure that all clinical tests are accurate and reliable.^22^ This is especially important as currently there is no gold standard serological reference method available for SARS-CoV-2.^23^ In developing a reliable antibody testing strategy, the focus is often placed on performance characteristics such as sensitivity and specificity, with operational characteristics, such as ease of use, test type, specimen type and time to results often deemed less important factors. A balance between these two elements is vital for the widespread endorsement of any serological testing method.^24^

Many point-of-care (POC) lateral flow immunoassays (LFIA) for the detection of SARS-CoV-2 antibodies in blood also exist. However, serious questions have been raised around the specificity, sensitivity, reproducibility and supply of such tests.^23,24,25^ Conversely though, their speed, low cost and relative ease of use may have applicability in screening for large populations, provided performance characteristics can be proven via clinical validation. The WHO Coordinated Global Research Roadmap for SARS-CoV-2 defines the requirement for simplified, rapid POC testing technologies that allow infection screening to be decentralised from the laboratory to remote patient management sites and operated by general healthcare staff without the requirement for specialised training.^8^

ELISA-based tests for detection of SARS-CoV-2 offer an alternative to the POC tests. Although more complex than rapid POC assays, ELISA are more sensitive (PPA) and specific (NPA) and can be performed in a high-throughput manner.^9,26^ Closed platforms such as the Abbott SARS-CoV-2 IgG assay, a chemiluminescent microparticle immunoassay (CMIA), and the Roche Elecsys^®^ Anti-SARS-CoV-2 antibody tests are designed to run on specialised equipment (Abbott’s ARCHITECT™ i1000SR and i2000SR; Roche’s Cobas™ e-analyser) in hospitals and reference laboratories by highly trained technicians. However, the limitation of closed platform systems from a single supplier is around supply of reagents, analogous to the problems with reagents for PCR testing.^27^

The clinical performance of the Trinity Biotech Captia™ SARS-CoV-2-IgG kit in a healthcare worker population was evaluated as part of the CAST study. The Positive Percent Agreement (or estimated sensitivity) and the Negative Percent Agreement (or estimated specificity) were 95.92% and 100% respectively at >14 days post swab, providing a reliable and robust methodology for determining IgG levels in healthcare workers with impactful utility in the medium to long term management of the COVID-19 pandemic.

There has been much discussion regarding antibody levels and their potential associated protection against SARS-CoV-2 re-infection. A study on SARS, a similar coronavirus to SARS-CoV-2, showed recovered individuals maintained neutralising antibodies for two years on average.^28^ In addition, antibody responses in individuals with laboratory-confirmed MERS-CoV infection lasted for at least 34 months after the outbreak.^29^ Long *et al*. and more recently Ward *et al*. have suggested a rapid decay of anti-SARS-CoV-2 IgG, although our findings do not support this assertion.^30,31^ Ward’s seroprevalence study on a large population in the U.K using lateral flow immunoassay testing found that healthcare workers along with ethnic minorities and care home workers were disproportionately affected by the pandemic with more positive test results.^32^ Interestingly, higher numbers of healthcare workers demonstrated persistence of antibody levels over serial time points whereas an overall decline in the general population was observed in this study.^29^

We report that over 90% of our study participants had detectable levels of anti-SARS-CoV2 IgG in their serum up to six months post infection. In our laboratory-confirmed positive participants, 98% were seropositive using the Captia™ assay (Trinity Biotech) at day 50 and 100, with 91% seropositive at day 160+, indicating a small decline in the number of participants maintaining a seropositive status. When using the Architect™ assay (Abbott) on the same participant cohort, only 80% of participants were detected as being seropositive at day 100. The Captia™ assay (Trinity Biotech) utilises the Spike protein as a capture antigen while the Architect™ assay (Abbott) utilises the Nucleocapsid protein, which is interesting to note as the Abbott assay is a serological assay utilised by the Irish Health Service Executive (HSE) for their seroprevalence study.^18^

Recommendations for healthcare workers at the time of the study were to adhere to strict public health guidelines; namely use of appropriate Personal Protective Equipment (PPE), social distancing where possible, careful hand hygiene and appropriate cough etiquette. A series of additional measures were introduced to the hospital at various points between March and July 2020 (Figure 2). These protective measures helped to reduce infection levels within the hospital. With 55% of positive cases reporting additional household members being symptomatic, household/community transmission rather than workplace transmission became the most common route of infection transmission for our hospital staff. High local hygiene standards and introduction of additional hygiene measures led to a rapid decrease in numbers of staff infections. Of the 54 positive staff diagnosed during the study timeframe, 45 close contacts were identified with 62% of these close contacts (28/45) becoming symptomatic and requiring subsequent testing. However only six of the close contacts identified returned a positive result (13%), with two thirds of cases linked to household or community transmission.

More recent reports across media platforms and in the literature describe long term or ongoing symptoms of the virus which greatly impacts the physical and mental health of those affected.^5,6,7^ This aligns with our findings whereby post-viral symptoms persist in 50% of our cohort of COVID-19 positive participants. The true ongoing health consequences among HCWs have yet to be realised. Early identification of SARS-CoV-2 positive HCW is crucial to avoid virus transmission between staff members and to maximize the available workforce.

### Study Limitations

This study was carried out shortly after the first pandemic peak in March 2020. Although our study cohort reported mild symptoms with few hospitalisations, as healthcare workers ongoing contact with each other and with patients increased the overall risk category of these individuals. Therefore, a limitation of this study is that our study group is a high risk group compared to the general population.^29^

## Conclusion

Accurate testing with high sensitivity and specificity is essential in helping understand and manage the SARS-CoV-2 pandemic. Healthcare workers are among the high risk groups in society with higher numbers of positive tests amongst the population. Longevity of antibody status as well as transmission of the virus and ongoing symptoms in this population is of great interest. Our findings confirm positive antibody status in 91% of those with confirmed PCR tests up to 160+ Days post infection. The Captia™ SARS-CoV-2 IgG kit provides a methodology for detecting IgG antibodies in healthcare workers and other populations with impactful utility in the medium to long term management of the COVID-19 pandemic.

## Data Availability

Data is available on a secure password protected database at the Rotunda Hospital

## Acknowledgements

We would like to acknowledge and thank the staff of the Rotunda Hospital who participated in this study. We would also like to thank the following departments and staff, who contributed to recruitment, sample collection, data entry and study support: Laboratory Department, Occupational Health Department, Phlebotomy Department, Swabbing Team, Contact Tracing Team and the Rotunda/RCSI Research Department.

## Study Contribution

Study Conception and Design – JG, ET, FC

Protocol Development -JG, ET, FC, KE, KM, DN, FM

Patient Recruitment and testing - JG, ET, FC

Data Entry – RC, RML, SB

Viral Testing – DLB, JOL, RD

Serological Analysis - KE, KM, DLB, JOL

Data Analysis - JG, ET, FC, KE, KM,

Manuscript Preparation - JG, ET, FC, KE, KM, DN, FM, JOL, DLB, SB, RC, RML

All authors approve of the final manuscript

